# Is genetic diversity of *Plasmodium falciparum* suggestive of malaria endemic zone in West Africa? An attempt to respond by a cross-sectional comparative study between Senegal and Nigeria

**DOI:** 10.1101/2020.08.27.20183178

**Authors:** Mary M. Oboh, Tolla Ndiaye, Khadim Diongue, Yaye D. Ndiaye, Mouhamad Sy, Awa B. Deme, Sarah K. Volkman, Aida S. Badiane, Alfred Amambua-Ngwa, Daouda Ndiaye

## Abstract

**Background:** Characterization of malaria parasite populations in different endemic settings (from low to high) could be helpful for ascertaining the progress of malaria interventions in endemic settings. The present study aims to compare *Plasmodium falciparum* parasite population structure between two West African countries with very different level of endemicity using *P. falciparum* allelic polymorphic markers: msp1 and msp2.

**Methods:** Parasite genomic DNA was extracted from 187 dried blood spot collected from confirmed uncomplicated *P. falciparum* malaria infected patients in Senegal (94) being at the pre-elimination stage in most part of the country and Nigeria (93) which is still at the control stage. Allelic polymorphism of msp1 and msp2 genes were assessed by nested PCR.

**Results:** In Senegal as well as in Nigeria, K1 and IC3D7 allelic families were the most represented for *msp1* and *msp2* genes respectively. A higher multiplicity of infection (MOI) was found in both study sites in Senegal (Thies:1.51/2.53; Kedougou:2.2/2.0 for *msp* 1/2) than from sites in Nigeria (Gbagada: 1.39/1.96; Oredo: 1.35/1.75]). The heterozygosity of *msp 1* gene was higher in *P. falciparum* isolates from Senegal (Thies: 0.62; Kedougou: 0.53) than isolates from Nigeria (Gbagada: 0.55; Oredo: 0.50). In Senegal, K1 alleles were associated with heavy (28) than with moderate (18) infections, in Nigeria however, equal proportions of K1 were observed in both infection types. The IC3D7 subtype allele of the *msp 2* family showed high occurrence in heavily infected individuals from both countries (Senegal- 32; Nigeria- 26) than in the moderately infected participants.

**Conclusion:** With the unusual high genetic diversity obtained in low endemic setting in Senegal and low genetic diversity in a high endemic Nigerian setting, multiple holistic approach should be employed in evaluating the actual transmission of a place in order to effectively direct control measures.

## Introduction

Malaria, a vector-borne infection continues to ravage public health havoc in many endemic countries in tropical and sub-tropical parts of the world [1]. Though a very preventable disease, malaria accounted for 228 million and 405 000 cases and death respectively in 2018 alone [2], A substantial gain in relative comparison with cases and fatality of 2010 and 2015 respectively has been recorded. This progress made so far is on a plateau where significant improvement cannot be clearly distinguished. In 2018 alone, malaria control and elimination efforts received a massive USD 2.7 billion in investment, nearly three quarters of which was targeted towards the African region. Regardless of this, the region contributed enormously to malaria cases (93% in 2018) and mortality (94%) within the same time period [2].

Malaria in Africa, majorly cause by *Plasmodium falciparum* continues to be a serious and debilitating infection with almost all age group at risk of the disease in endemic areas. Senegal and Nigeria are two countries located in western part of Africa with varied levels of interventions, endemicity and transmission. In Senegal, all population are exposed to malaria and transmission increases gradually from the northern to the Southern part. This is very much correlated with the hyperendemic and hypoendemic nature observed in the south (annual incidence is greater than 100/1000 inhabitants) and North (annual incidence rates are now less than 5/1000 inhabitants) respectively [3] In 2017, malaria accounts for 395 706 cases, 284 deaths and by 33.45 % among children under 5 were estimated in Senegal [4]even so malaria control has been intensified more than ten years. The endemicity of malaria in Nigeria varies between the six geo-political zones of the country, with south west having a mix of meso- and hypo-endemic (1-50%) situation, the south south, south east, north east and north central uniformly hypo-endemic (20-39%), while the north western part is highly meso-endemic (40-49%). Annually, 40% of Nigeria gross domestic product is spent on malaria control [5], yet the situation still look gloomy.

Of the several factors contributing to this persistent health predicament, mis-diagnosis as a result of the inability of the point-of-care rapid diagnostic tool (RDT) to pick up infections with histidine-rich protein 2 and 3 (HRP2 and HRP3) deletion, development of resistance by the parasite and vector to antimalarial chemotherapy and insecticide respectively [6], all take the front burner.

Hence, the development of an effective malaria vaccine that can be readily available to at risk individuals in endemic areas cannot be overestimated. Nevertheless, this initiative is being impeded by the enormous parasite diversity [7-9] which renders the vaccine almost ineffective. In moderate and high transmission areas such as in Africa, the probability of a person being infected with numerous parasite clones at the same time is very high [10-14], These clones/strains are often a reflection of the transmission intensity or endemicity level of an area and as such, impacts the immune system of residing persons. Ultimately, the interplay between parasite clones and the immune selective pressure has a profound impact on the success of any effective vaccine developed.

Although, the only approved malaria vaccine (RTS, S) gives a 30% protection, albeit to state that, multiple booster doses are required to achieve that level of defense, the development of an efficacious vaccine becomes imperative. In addition, the pathogen genome diversity is known to impact intervention strategies including drugs and insecticides, characterization of malaria parasite populations in different endemic settings (from low to high), becomes much more needed to help in the development of a second-generation vaccine.

The asexual blood stage antigens- merozoite protein 1 and 2 (MSP 1 and MSP 2) along with the glutamate-rich protein (GLURP) are highly diverse antigens being exploited for vaccine advancement [15, 16]. Furthermore, they have been used in various studies in evaluating the different circulating clones of *P. falciparum* and/or determining the impact of intervention studies [17, 18] in order to ascertain the progress of such interventions.

There is paucity of data on the comparative study between *P. falciparum* parasite population structure among West African countries which present very different level of endemicity. Therefore, the objective of this study was to compare genetic diversity of *P. falciparum* in the *msp* 1 and 2 genes from Nigeria which is still at the control stage and Senegal being at the pre-elimination stage in most part of the country, using length polymorphism of genotype parasite population. A secondary objectives was to determine the multiplicity of infection and heterozygosity, both of which reflect the transmission intensity as affected by intervention.

## Materials and Methods

### Ethics consideration

The study was ethically approved by the Institutional Review Board of the Nigerian Institute of Medical Research, Lagos and the Senegalese National Ethics committee. Written informed consent and/or assent was obtained from the parents and guardian of children prior to recruitment.

### Study sites and samples collection

This study was carried out in two West African countries: Nigeria and Senegal.

In Nigeria, isolates were collected in two Local Government Areas of Lagos states namely Kosofe (O6°28⍰N 003°22⍰E) and Ikorodu (O6°33⍰N 003°35⍰E). Description of study area has been done in our earlier study [19]. Briefly, Lagos state shares a border with the Republic of Benin and is hypo-endemic in most part with a 1.9% prevalence rate in children age 6–59 months [20]. This is in part due to the expansion of insecticide-treated nets (ITNs) coverage, occurrence of indoor residual spraying (IRS) in many of its LGAs [21],

In Senegal, samples were collected in two areas with different endemicity levels, Kedougou located in the south-east and Thiès in western Senegal. In Kedougou, malaria transmission is seasonal from July to December, with an entomological inoculation rate (EIR) of 20 to 100 infectious bites/person/ year and an incidence higher than fifteen malaria cases per 1,000 habitants. While in Thies, the malaria situation is hypoendemic, with an average 0-20 infectious mosquito bite annually and an incidence of five to fifteen malaria cases per 1,000 habitants. The malaria seasonal transmission in this area coincides with rainy season which generally last up to four months (September to December) [22, 23].

Febrile patients visiting health facilities in these areas during malaria transmission season were recruited in this study. Blood samples were collected on filter-paper from patients who met the following inclusion criteria: residence 15-km radius of health facilities, presence of febrile condition (axillary temperature ≥ 37.5 °C) in the previous 48 h, age ranging from 6 months to 75 years and uncomplicated *P. falciparum* malaria with parasite density ≥ 1000 asexual forms per microliter. Patients who presented signs or symptoms of severe malaria as defined by World Health Organization (WHO) [2] and pregnant women were not included.

### Parasite DNA extraction and allelic typing of *P. falciparum msp1* and *msp2* genes

Parasite genomic deoxyribonucleic acid (gDNA) was extracted from filter paper using QIAamp DNA Mini kit (QIAGEN, USA) according to the manufacturer’s instructions.

The polymorphic loci of *P. falciparum msp1* block 2 (K1, MAD20 and RO33) and *msp2* central region (IC3D7 and FC27) were used as genetic markers for genotyping of parasite populations following a previously described nested PCR protocol [24, 25] (supplementary **Table 1)**. All PCR reactions were carried out in a final volume of 20 μl containing: 1 μl of gDNA, 6 μl GoTaq Green Master Mix (Promega), 1 μl (0.5 μM) of each primer, and 11 μl nuclease free water. In both rounds of reaction, 1 μl of gDNA and PCR amplicon were used respectively as templates for nest 1 and 2 amplifications.

Cycling conditions for both PCR were as follows: initial denaturation at 95 °C for 5 min, followed by 35 cycles of denaturation at 94 °C for 1 min, annealing at 58 °C for 2 min (61 °C for 2 min for the nested reaction) and extension at 72 °C for 2 min; a final extension was carried out at 72°C for 3 min. Positives (3D7 and Dd2) and negative (DNase free water) controls were systematically incorporated in each PCR run.

The nested PCR product were resolved in 2% agarose gels electrophoresis stained with ethidium bromide and visualized under UV trans-illumination (VersaDoc®, Bio-Rad, Hercules, USA). The sizes of PCR fragments were estimated using 100 bp molecular weight ladder (Maker). Presence of more than one genotype was taken as a polyclonal infection, while a single allele was considered as a monoclonal infection. If fragment sizes were within a 20 bp interval, alleles in each family were considered the same [26].

### Statistical analysis

The online Biostatgv was used for statistical analysis. Isolates presenting more than one allele were considered multiclonal isolates. The chi-square test (X^2^) was used to compare the frequencies of multiclonal isolates between localities and countries.

The mean multiplicity of infection (MOI) was calculated as the quotient of the total number of genotypes for each marker and the number of PCR-positive samples. Thus, MOI was calculated by dividing the total number of alleles detected for *msp1* and *msp*2 genes by the total number of samples [27],Student’s t test was used to compare MOI between localities. We used the expected heterozygosity (He, which is a measure of genetic diversity) and genetic differentiation (F_ST_) to assess population structure of parasites. Heterozygosity was calculated using the following formula He = [n/(n-1)] [(1-ΣPi2)], where n = sample size, Pi = allele frequency as described by [28]. A p-value of ≤ 0.05 was considered suggestive of a statistically significant difference.

## Results

A total of 187 participants were recruited for the study with almost equal proportion from the two countries (94 from Senegal and 93 from Nigeria). The mean age was not different from each country and locality within the countries. Although, participants were randomly enrolled into the study, however, more males (108) partook in the study than females (79) **(Table 1)**.

**Table 1:** Characteristics of the study participants in four different endemic sites located in the two West African countries.

|  | Senegal |  | Nigeria |  |
| --- | --- | --- | --- | --- |
|  | Thies | Kedougou | Gbagada | Oredo |
| Number (%) | 47 (50) | 47 (50) | 29 (31.2) | 64 (68.8) |
| Age |  |  |  |  |
| Mean | 26.8 | 20.4 | 24 | 27.8 |
| Range | [9-72] | [2-67] | [5-65] | [1-70] |
| Sex |  |  |  |  |
| Male | 41 (87.2) | 23 (48.9) | 16 (55.2) | 28 (43.8) |
| Female | 6 (12.8) | 24 (51.1) | 13 (44.8) | 36 (56.2) |

### Genetic polymorphism of msp1 and msp2 genes in Senegal and Nigeria

For *msp1* gene, findings from the two countries practically were overlaid. K1 allelic family was predominant in Senegal and Nigeria with frequencies of 44% and 48%, respectively. RO33 allelic family was less represented in the both country with the frequency of 6% in Senegal and 15% in Nigeria. With regards to multiple allelic family infections, K1/RO33 combination was the most predominant in both countries, 17% in Senegal and 7% in Nigeria. However, the K1/MAD20/RO33 trimorphic allelic infections was less found in Senegal (3%) and Nigeria (2%).

The fragment size of K1 allele type observed in Senegal (100-350bp) was slightly different from that seen in *P. falciparum* isolates from Nigeria (190-300bp). Similar pattern was also noticed with the MAD20 alleles where sizes in Senegal ranges from 100-300 bp while those from Nigeria ranges from 200-300 bp. In addition, a different pattern was observed in RO33 where only one clone type was seen in Nigeria (160 bp) as against observation of multiple clones in Senegal (160-240bp) (Fig. 1A and Fig. 1B).

Regarding *msp1* gene, there was no observable difference in the frequency of IC3D7 allele in Senegal (57%) and Nigeria (58%), while the FC27 frequency was higher in Senegal (26 %) than in Nigeria (16%). Similar pattern was noticed with the IC3D7/FC27 dimorphic allelic family in both countries. For the allelic size polymorphism, the size of the IC3D7 clone of *msp2* ranges from 300-700bp in Senegal and 300-800bp in Nigerian *P. falciparum* isolates while that of the FC27 showed varied sizes from 200-600 bp in Senegal and 290-550 bp in Nigeria (Fig.1C and Fig.1D).

**Figure 1:**
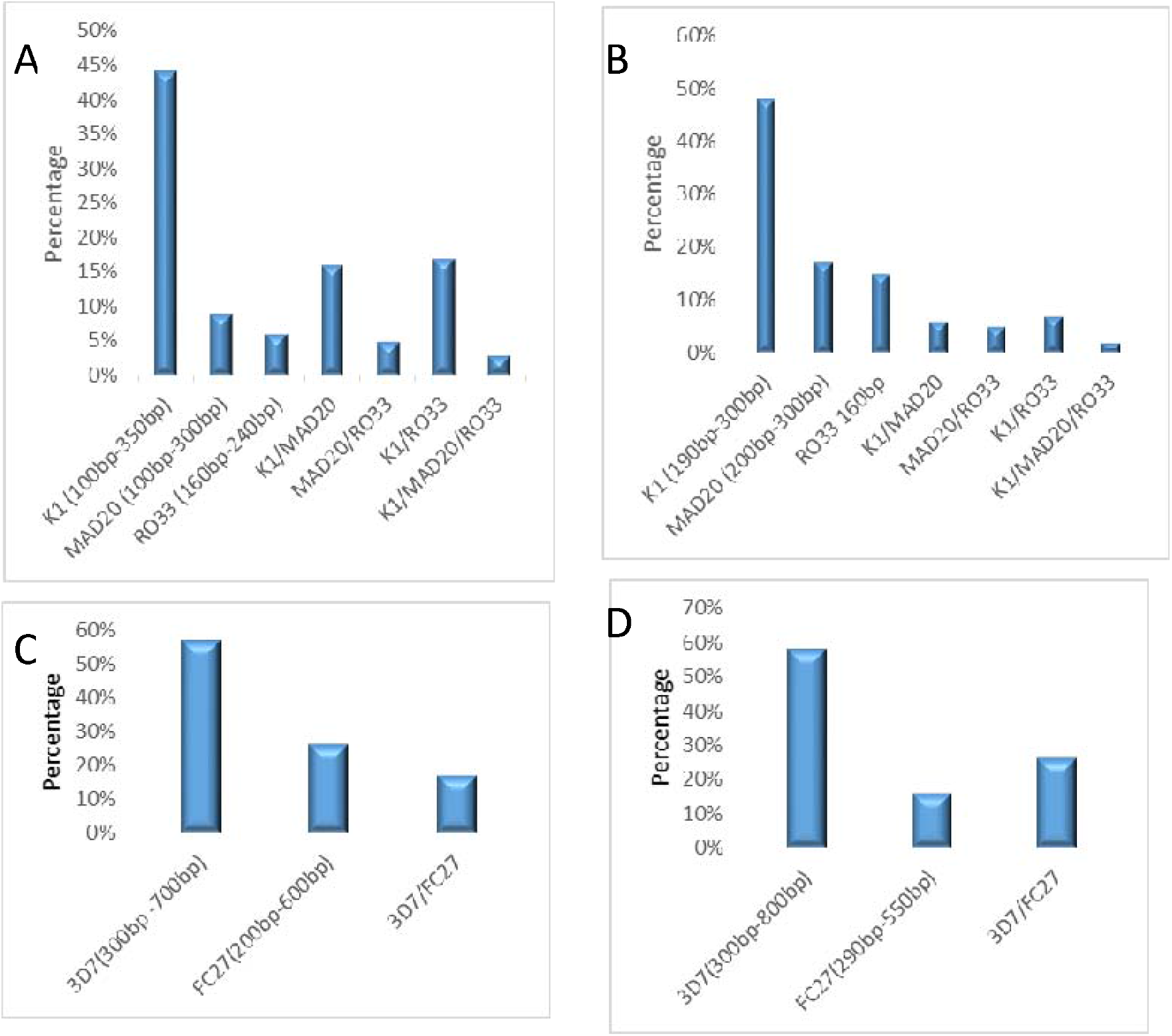
Genetic polymorphism of allele fragment sizes of *P. falciparum msp 1* in A) Senegal B) Nigeria and *msp 2* in C) Senegal D) Nigeria

### Multiplicity of *P. falciparum* infection and heterozygosity of *msp1* and *msp2*

The presence of multiple clones of infection, otherwise known as MOI was consistently higher in both study sites in Senegal (Thies, 1.51/2.53; Kedougou, 2.2/2.0 for *msp* 1/2) than the sites in Nigeria (Gbagada, 1.39/1.96; Oredo, 1.35/1.75). Consequently, the heterozygosity of *msp* 1 gene was higher in *P. falciparum* isolates from Senegal (Thies, 0.62; Kedougou, 0.53) than isolates from Nigeria (Gbagada, 0.55; Oredo, 0.50). However, the heterozygosity of *msp* 2 gene was not different for the two countries **(Table 2)**.

**Table 2:** Multiplicity of infection and heterozygosity of msp1 and *msp*2 of *P. falciparum* from Senegal and Nigeria

| Gene |  | Senegal |  | Nigeria |  | p-value |
| --- | --- | --- | --- | --- | --- | --- |
|  |  | Thies | Kedougou | Gbagada | Oredo |  |
| *msp 1* | MOI | 1.51 | 2.2 | 1.39 | 1.35 | 0.00* |
|  | He | 0.62 | 0.53 | 0.55 | 0.50 | 0.89 |
| *msp 2* |  |  |  |  |  |  |
|  | MOI | 2.53 | 2.00 | 1.96 | 1.75 | 0.39 |
|  | He | 0.48 | 0.44 | 0.47 | 0.48 | 0.77 |
|  | MOI (both genes) | 2.65 | 2.68 | 1.64 | 1.57 | 0.72 |

MOI- multiplicity of infection, He- heterozygosity. \*P-value less than 0.05 significant

### Genetic polymorphism of msp1 and msp2 genes by age range

Attempts to visualise the genetic diversity of various allele types in both msp1 and 2 in individuals infected with different parasite density showed some level of stratification. For msp1 allele type, K1, MAD20, RO33 and their various allele combinations were absent in individuals with light infections (50-499 parasites/ μl) from both countries. There was more observation of K1 in individuals with heavy infections (28) than those with moderate infections (18) from Senegal. However, in Nigeria, equal proportions of K1 were present in both moderate and heavy infections. Individuals with heavy infections from Senegal showed the highest proportion of K1/MAD20 (14) allele combination and same pattern was observed for the K1/RO33 mixture (13).

The IC3D7 subtype allele of the msp 2 family showed high occurrence in heavily infected individuals from both countries (Senegal, 32; Nigeria, 26) than in the moderately infected participants. Similar pattern was noticed with the FC27 allele **(Figure 2a and b)**.

**Figure 2a:**
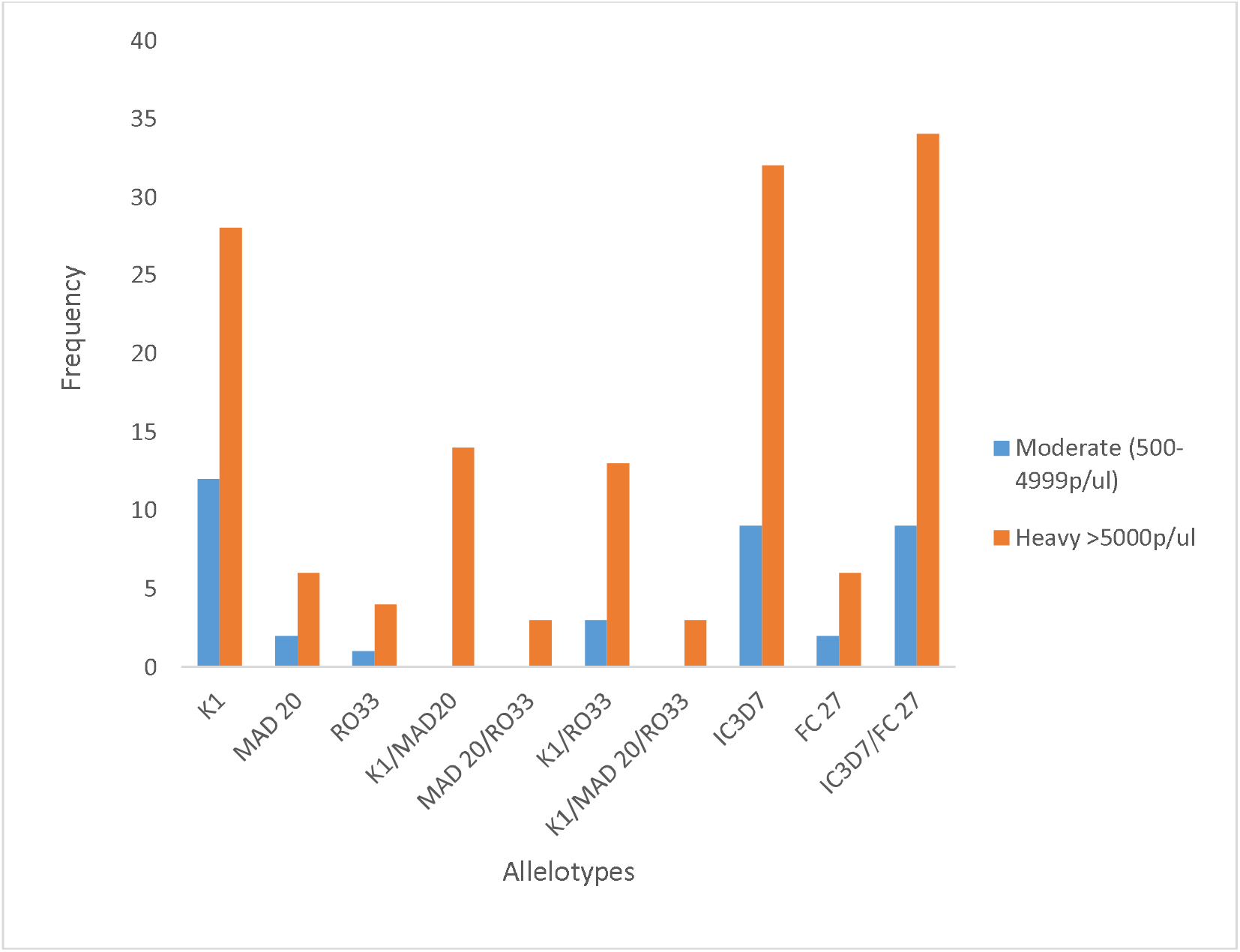
Genetic diversity of *P. falciparum msp*1 and *msp*2 stratified by parasite density in infected individuals from Senegal

**Figure 2b:**
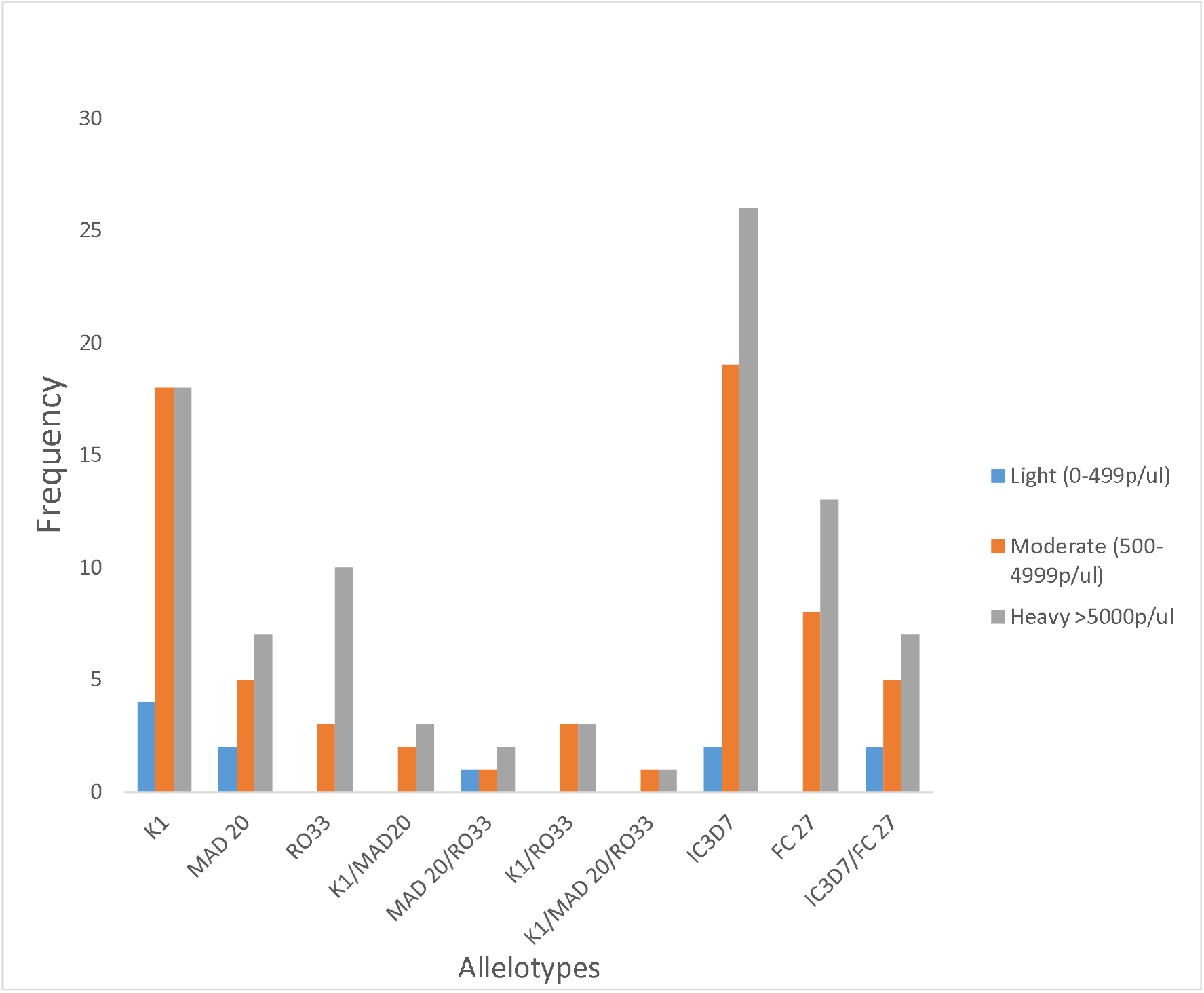
Genetic diversity of *P. falciparum msp*1 and *msp*2 stratified by parasite density in infected individuals from Nigeria

## Discussion

Control interventions targeting malaria parasite continues to face multiples hurdles from development of drug resistance, to insecticidal resistance by the vector, unavailability of a reliable vaccine to confer the necessary protection against the parasite. Hence, there is a critical need for continuous monitoring and evaluation of the effectiveness of control measures. To this end, we have attempted to evaluate the diversity of *P. falciparum* merozoites surface protein 1 and 2 from two West African countries with significantly different endemicity. Generally, a hypo-endemic area on one hand (Senegal) and a meso- hyper endemic area on the other hand (Nigeria).

Multiplicity of infection (MOI) an indication of multiclonal isolates and usually associated with the level of malaria transmission [29-31] was evaluated in this study. High MOI for both the *msp* 1 and 2 genes was observed for both study locations in Senegal as against the moderate MOI seen in Nigeria. Similar results were reported by Niang et al, in 2017 (Ref). This is an atypical situation as overall, Senegal is categorized as a country under the pre-elimination stage and as such is supposed to show a moderate to low MOI while Nigeria has intense transmission going on in all regions of the country. The reason for this unexpected finding could be attributed to dynamic transmission indices in different foci of a malaria endemic country.

Heterozygosity which is a measure of the genetic diversity of the two genes showed *Pfmsp1* from Senegal to be more diverse than that from Nigeria, however, same level of diversity for *Pfmsp2* was observed in both countries. Again, the reason for this reversed pattern (i.e high diversity seen in low transmission settings in Senegal and low diversity seen in high transmission setting in Nigeria) is unknown. Moreover, there was a difference in the clonal structures of all allelic families (K1, MAD20, 3D7 and FC27) from both countries with Senegal showing the more sub-structured *Pfmsp1* and *Pfmsp1* populations. Nevertheless, similar trend has been observed in Kingdom of Eswatini where a high genetic diversity was obtained in a low transmission area [32]. Perhaps, this result underscore the need for a more comprehensive evaluation of transmission using different epidemiological tools.

The high occurrence of *msp1* K1 allele in heavy and light infections from both countries is not knew as various studies [33] have shown this allele to be associated more with this grade of parasite density. Whether the high parasite density is driving the selection of K1 or vice versa is yet to be determined. The strong presence of this allelic families was reported by many studies carried out in West Africa [30, 34], western Uganda [35] and Iran [36]. Similarly, high frequency of IC3D7 allele type of *msp2* gene was also observed with both grades of parasite density, while no particular pattern was noticed with the FC27 allelotype. This finding is in agreement with the study of Hamid *et al*., in 2013 [37] where IC3D7 alleles was more predominant in moderate and heavily infected individuals.

Taken together, we therefore conclude that evidence driving selection of these observed allelotypes in moderate and heavy *P. falciparum* individuals should be evaluated in a bidirectional manner and a more holistic approach should be employed in determining the true epidemiological situation of any malaria endemic country as this will help in a more targeted control measure

## Data Availability

All data generated from this study have been included in the manuscript

